# SARS-CoV-2 Antigen Tests Predict Infectivity Based on Viral Culture: Comparison of Antigen, PCR Viral Load, and Viral Culture Testing on a Large Sample Cohort

**DOI:** 10.1101/2021.12.22.21268274

**Authors:** James E. Kirby, Stefan Riedel, Sanjucta Dutta, Ramy Arnaout, Annie Cheng, Sarah Ditelberg, Donald J. Hamel, Charlotte A. Chang, Phyllis J. Kanki

## Abstract

The relationship of SARS-CoV-2 antigen testing results, viral load, and viral culture detection remains to be fully defined. Presumptively, viral culture can provide a surrogate measure for infectivity of sampled individuals, and thereby inform how and where to most appropriately deploy available diagnostic testing modalities. We therefore determined the relationship of antigen testing results from three lateral flow and one microfluidics assay to viral culture performed in parallel in 181 nasopharyngeal swab samples positive for SARS-CoV-2. Sample viral loads, determined by RT-qPCR, were distributed across the range of viral load values observed in our testing population. We found that antigen tests were predictive of viral culture positivity, with the LumiraDx method showing enhanced sensitivity (90%; 95% confidence interval (95% CI) 83-94%) compared with the BD Veritor (74%, 95% *C*I 65-81%), CareStart (74%, 95% CI 65-81%) and Oscar Corona (74%, 95% CI 65-82%) lateral flow antigen tests. Antigen and viral culture positivity were also highly correlated with sample viral load, with areas under the receiver-operator characteristic curves (ROCs) of 0.94-0.97 and 0.92, respectively. In particular, a viral load threshold of 100,000 copies/mL was 95% sensitive (95% CI, 90-98%) and 72% specific (95% CI, 60-81%) for predicting viral culture positivity. Taken together, the detection of SARS-CoV-2 antigen identified highly infectious individuals, some of whom may harbor 10,000-fold more virus in their samples than those with any detectable infectious virus. As such, our data support use of antigen testing in defining infectivity status at the time of sampling.

## Introduction

Ongoing management of the SARS-CoV-2 pandemic will require judicious use of available diagnostic testing modalities. Testing may be used in two general contexts, either for diagnosis of symptomatic illness or to identify asymptomatic carriers so that appropriate isolation precautions can be instituted and thereby mitigate the infectious risk to others.

There are two major SARS-CoV-2 diagnostic technologies: (1) nucleic acid amplification tests (NAATs), typically real-time polymerase chain reaction tests (PCR), and (2) antigen tests, which generally detect the presence of viral nucleocapsid (N) antigen. For the purposes of discussion, we will use PCR to represent all types of NAAT tests, recognizing that there are several widely used alternative detection technologies such as transcription mediated amplification. PCR tests are highly sensitive, and generally target conserved regions in more than one viral gene to ensure robustness even in the context of ongoing evolution of genetic variants. However, they are expensive compared to antigen tests and have slower turn-around time (TAT) when performed on large automated diagnostic platforms, which are usually located in central laboratories. Most antigen tests currently on the market are based on lateral flow immunoassay methods and detect the conserved N protein (1). In contrast to NAAT tests, antigen tests are fast, highly amenable to point-of-use, and inexpensive. Although antigen tests have lower sensitivity compared with PCR tests, it should be noted that antigen and PCR tests detect different components of the SARS-CoV-2 virus, neither of which may necessarily reflect live replicating virus.

With regard to appropriate selection and use of these alternative testing modalities, the ultimate question is how sensitive does a test need to be to serve specific goals of the pandemic response, given cost constraints and speed of testing requirements. A test with lower sensitivity may offer compelling benefit if it were able to reliably identify individuals who present an immediate infectious risk to others. Arguably, more frequent and repetitive testing of individuals using a test with lower sensitivity, may provide equivalent utility to more sensitive NAAT technology at lower cost and greater convenience (2).

An individual’s infectivity presumptively may be approximated by the infectivity of a diagnostic sample. Diagnostic samples may contain viable virus, non-viable/non-replicating virus components, and/or free viral nucleic acid. Antigen and PCR tests target different components of the SARS-CoV-2 virus, and detection by either technology does not necessarily reflect the presence of replication-competent, infectious virus and therefore infectiousness. However, viable virus can be cultured, i.e., it will infect and replicate in tissue culture cells. Therefore, tissue culture assays can provide an assessment of the presence and amount of viable virus, and, by inference, the infectivity of individuals from whom samples were obtained.

Therefore, we sought to compare the results of antigen and quantitative PCR viral load determination with viral culture detection of SARS-CoV-2 to further inform decisions on best utilization of antigen and PCR testing.

## Results

In our study design, the same patient specimens were tested by reverse-transcription (RT)-qPCR; antigen detection using four different commercial methods, three with FDA Emergency Use Authorization; and viral culture for SARS-CoV-2. This avoided the need for six separate sample collections from each patient and the inherent variation introduced during collection of multiple specimens from the same individual (3). Of note, typically, the instructions for use (IFU) for antigen tests indicate that sample from a single nasal collection swab should be eluted directly in the test specific extraction buffer. Therefore, we estimate that samples used for antigen testing in our protocol were ∼17 to 18-fold more dilute (see materials and methods section) than they would have been had a separate, dedicated swab been used directly for these tests.

A total of 206 samples positive for SARS-CoV-2 by RT-qPCR using the Abbott M2000 or Abbott Alinity m platforms (limit of detection ∼100 genome copies/mL) were analyzed by LumiraDx (n= 206), BD Veritor (n=204), CareStart (n=201), and Oscar Corona (n=193) antigen tests and by viral culture (n=181). For all but thirteen samples, sufficient sample volume was available to test using all four antigen testing methods. Samples were preselected to span the distribution of viral loads observed during PCR testing from March to June, 2021.

We first compared categorical agreement between antigen test and viral culture results. Viral cultures were performed by adding sample to VeroE6 cells and cultured for 13-14 days. Viral culture positivity was scored both qualitatively and quantitatively using RT-qPCR to detect SARS-CoV-2 in viral culture supernatants as described in the materials and methods section. Two-by-two contingency tables, sensitivity, specificity, positive and negative predictive values for qualitative comparisons of each antigen testing method to viral culture are shown in Tables 1-4. Sensitivity was 90% for LumiraDx and 74% for the other methods, using viral culture positivity as the gold standard. Specificity was 70% for LumiraDx and 91-92% for the other antigen test methods. Negative predictive values for the LumiraDx were approximately 10% higher than for other methods.

**Table 1.**
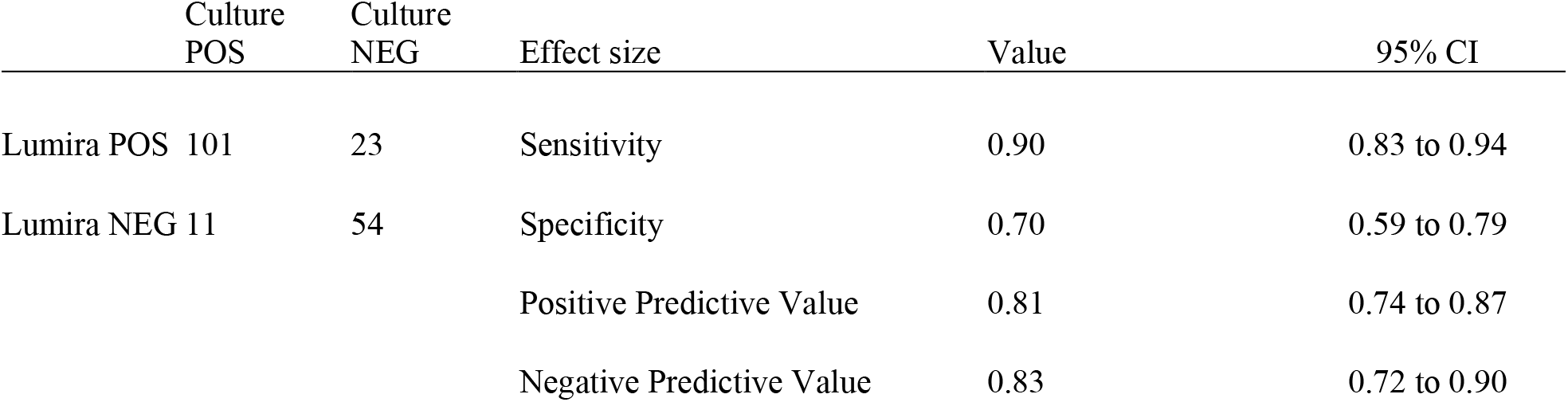
LumiraDx Antigen Results Versus Viral Culture.

**Table 2.** BD Veritor Antigen Results Versus Viral Culture.

|  | Culture<br>POS | Culture<br>NEG | Effect size | Value | 95% CI |
| --- | --- | --- | --- | --- | --- |
| BD POS | 82 | 6 | Sensitivity | 0.74 | 0.65 to 0.81 |
| BD NEG | 29 | 70 | Specificity | 0.92 | 0.84 to 0.96 |
|  |  |  | Positive Predictive Value | 0.93 | 0.86 to 0.97 |
|  |  |  | Negative Predictive Value | 0.71 | 0.61 to 0.79 |

**Table 3.**
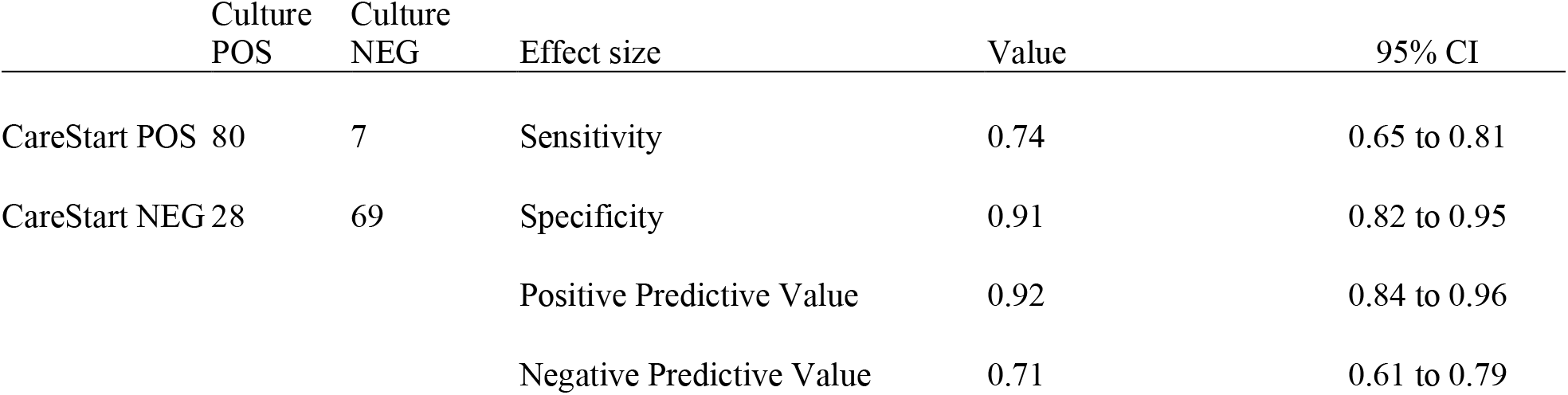
CareStart Antigen Results Versus Viral Culture.

**Table 4.**
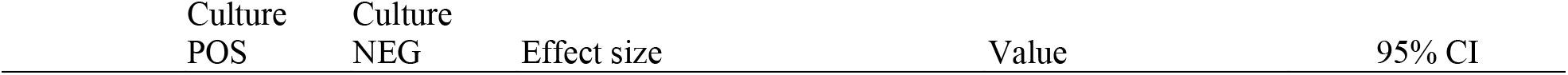
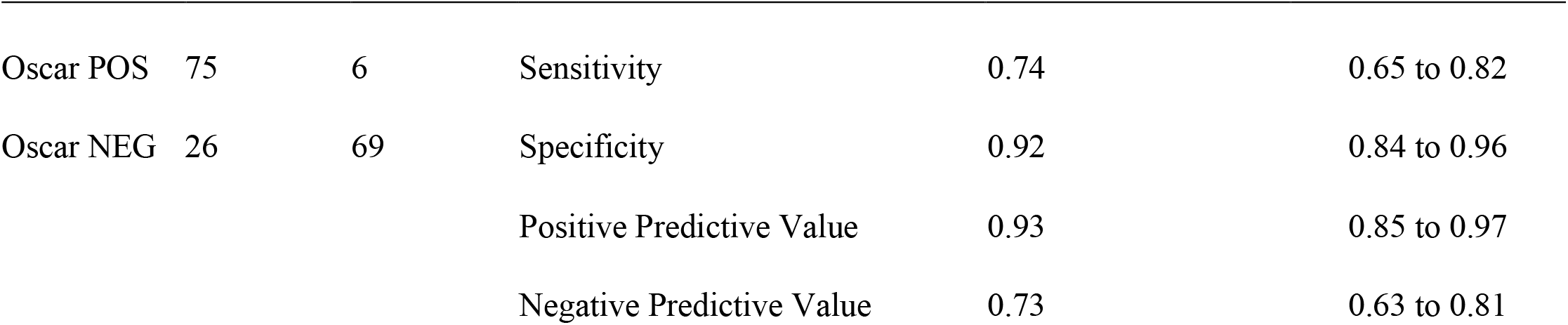
Oscar Corona Antigen Results Versus Viral Culture.

|  | Culture<br>POS | Culture<br>NEG | Effect size | Value | 95% CI |
| --- | --- | --- | --- | --- | --- |
| Oscar POS | 75 | 6 | Sensitivity | 0.74 | 0.65 to 0.82 |
| Oscar NEG | 26 | 69 | Specificity | 0.92 | 0.84 to 0.96 |
|  |  |  | Positive Predictive Value | 0.93 | 0.85 to 0.97 |
|  |  |  | Negative Predictive Value | 0.73 | 0.63 to 0.81 |

We then examined the quantitative relationship between sample viral load and the presence and quantity of culturable virus. We observed that levels of virus in day 3 viral culture supernatant were reasonably correlated (R^2^ = 0.55) with original sample viral load, with a sharp loss of detection of viable virus in samples with viral loads of less than ∼10^5^ genome copies/mL (Fig. 1).

**Figure 1.**
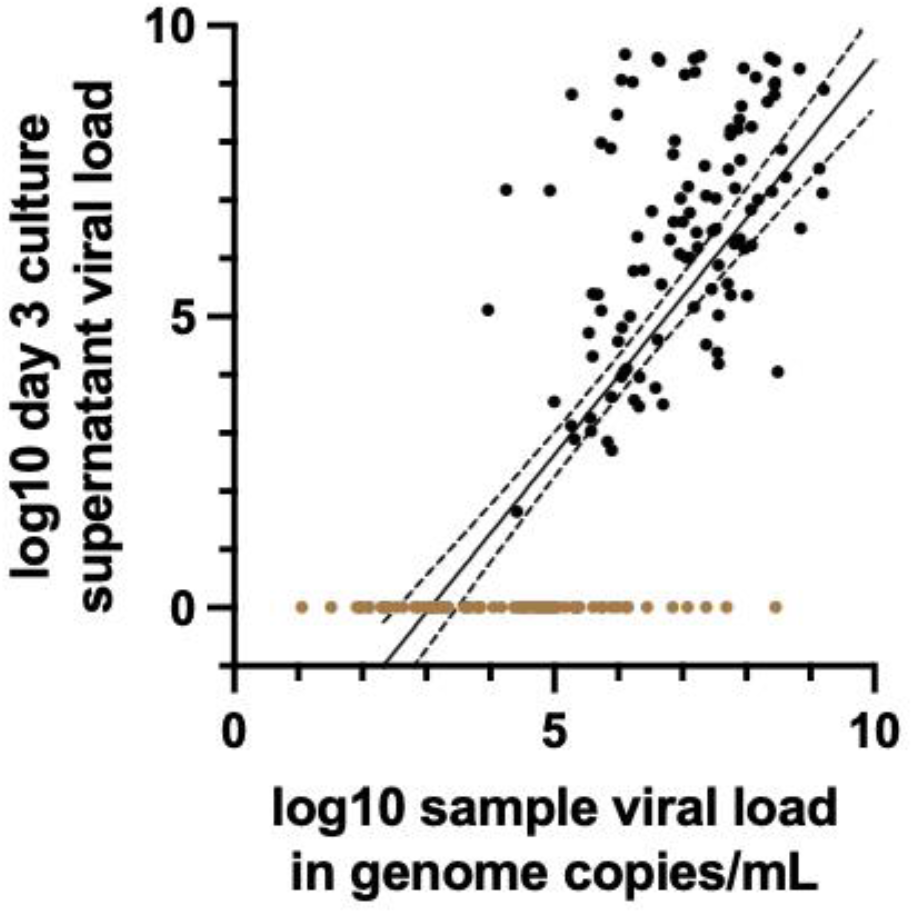
Quantitative Relationship Between Culturable Virus and Sample Viral Load. Day 3 viral culture supernatant from each cultured sample and its corresponding respiratory sample were each analyzed by RT-qPCR to determine respective viral loads (n=181). The viral load in log10 genome copies/mL of culture supernatant is plotted against the log10 viral load in genome copies/mL of the original patient sample. Linear regression (solid line) with 95% confidence intervals (dashed lines) is shown. R^2^ = 0.55. Samples with negative viral cultures (see materials and methods) for representation are assigned a y-axis, log10 value of 0 and are demarcated as colored brown dots.

Distributions of viral load results determined by RT-qPCR in samples, testing positive or negative, respectively, by respective antigen tests are shown in Fig. 2. Lateral flow assay (LFA)-based antigen tests detected samples with a viral load > ∼10^7^ genome copies/mL, with the exception of a small number of outliers. The microfluidic LumiraDx test detected samples with lower viral loads compared to the other antigen testing methods with a cutoff for consistent detection closer to 10^6^ genome copies/mL. Differences between log_10_ transformed mean viral loads of samples positive by LumiraDx and positive by each of the LFA methods, respectively, were statistically significant for all pairwise comparisons (adjusted *P* < 0.05), as were differences in (geometric) mean viral loads of samples testing negative by LumiraDx and negative by each of the LFA methods, respectively, using the Holm-Sidak’s multiple comparison test. However, BD Veritor, CareStart and Oscar Corona tests were indistinguishable in all pairwise comparisons with one another (adjusted *P* = 0.99).

**Figure 2.**
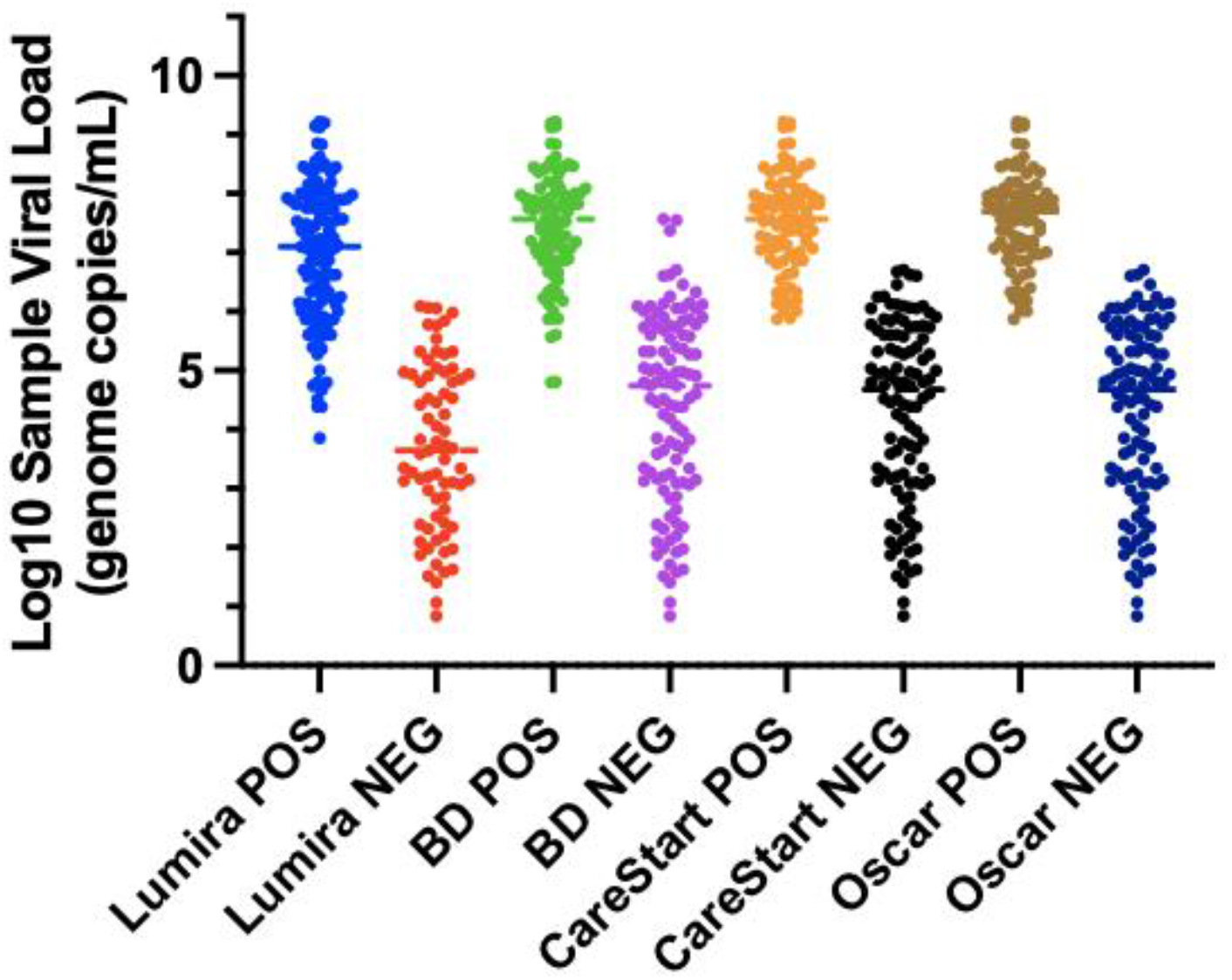
Antigen Testing Results Compared with Log10 Viral Load. Viral load in log10 genome copies/mL. POS = positive antigen test result. NEG = negative antigen test result. Lumira = LumiraDx antigen test; BD = BD Veritor antigen test; Oscar = Oscar Corona antigen test.

Receiver-operator characteristic curves (ROC) were plotted to determine the viral load cutoffs which would reasonably predict detection by viral culture and antigen testing, respectively (Fig. 3). Notably, a viral load cutoff of ∼10^5^ was highly sensitive for predicting a positive viral culture without undue loss of specificity. A viral load cutoff of ∼10^4^-10^5^ was reasonably sensitive for predicting a positive LumiraDx result without undue loss of specificity. Viral load cutoffs of 10^5^-10^6^ likewise were reasonably predictive of positive BD, Oscar and CareStart test results. The ROC area under the curve (AUC) was > 0.94 for all comparisons (Fig. 3B-E), indicating that viral loads could serve as a reasonable surrogate for predicting the presence of culturable infectious virus and detectable antigen (and vice versa). Notably, viral load cutoffs for detecting positive viral cultures and positive antigen tests were similar, further supporting similar qualitative detection by viral culture and antigen tests.

**Figure 3.**
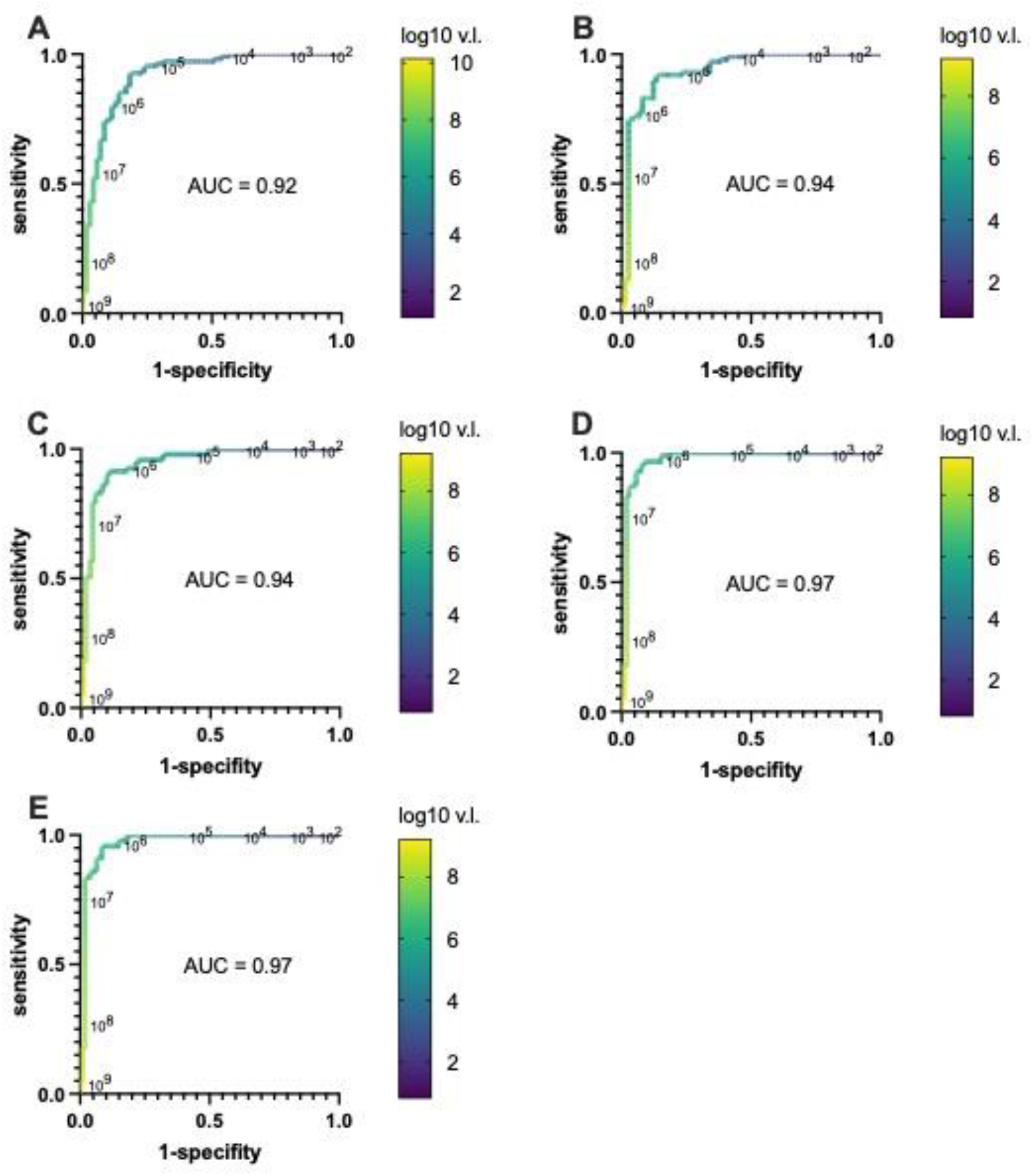
Receiver operator characteristic curves (ROC) comparing SARS-CoV-2 sample viral load levels as a predictor of viral culture and antigen detection. For each plot, sensitivity versus 1-specificity was plotted for each viral load value (genome/copies/mL) determined by RT-qPCR for each sample in our study when used as a lower limit threshold for scoring positive and negative detection for all other viral load results with qualitative viral culture or antigen test determinations, respectively, as the comparators. (A) Log10 viral load (v.l.) in genome copies/mL versus detection by viral culture. (B) Log10 viral load versus LumiraDx antigen detection. (C) Log10 viral load versus BD Veritor antigen detection. (D) Log10 viral load versus Oscar Corona antigen detection. (E) Log10 viral load versus CareStart antigen detection. Viral load values along the ROC curves are labeled in log10 intervals and demarcated in color as indicated in accompany heatmap legend bar. AUC (area under the curve) for each ROC curve is denoted on respective plots.

Three hundred SARS-CoV-2 RT-qPCR negative samples were also tested by all four antigen tests. All antigen tests were negative, supporting high specificity of the antigen assays.

## Discussion

The evolving COVID-19 pandemic has brought a growing need for more and diverse diagnostic test methods. The detection of SARS-CoV-2 RNA by RT-qPCR is the gold standard for laboratory diagnosis of COVID-19, yet it is well recognized that the assay may detect RNA fragments or viral debris that do not correlate with viable infectious virus. For example, in the Syrian golden hamster model, transmissibility of SARS-CoV-2 correlated well with detection of infectious virus by culture, but not with positive RNA results by qPCR (4). Therefore, qPCR may identify a large number of patients who are infected, but who may not necessarily be infectious to others. SARS-CoV-2 viral culture may be a better surrogate of infectivity, but is currently impractical and rarely used as a primary testing modality due to its stringent biosafety requirements (Biosafety Level 3; BL3) requirements, assay complexity, and low throughput (5, 6).

Antigen test methods are an inexpensive and more rapid test method compared to most NAAT tests and are amenable to point-of-care settings, for example, use at schools and for self-testing. We therefore compared performance of antigen tests to PCR and viral culture to assess the ability of antigen tests to identify infected and infectious individuals. Our general findings were that antigen tests largely predicted ability to culture live virus. Furthermore, culture and antigen tests were both consistently positive at higher viral loads as determined by RT-qPCR and negative at lower viral loads.

Previous studies have also correlated viral culture with qPCR testing, describing positive cultures from upper respiratory tract samples between 6-9 days after infection (6-9). In our study, a viral load of cutoff of 100,000 copies/mL was 95% sensitive (95% CI, 90-98%) and 72% specific (95% CI, 60-81%) for predicting viral culture positivity. Other studies of SARS-CoV-2 culture have reported similar findings, where viable virus culture was described from samples with 250,000-1,000,000 copies/mL (9, 10) or expressed alternatively as cycle threshold (Ct) value cutoffs of 24-35 (11, 12). Note, we converted from Ct values to viral loads (in units of genome copies/mL), since Ct values cannot be compared from assay to assay (3).

Presumptively, individuals with the highest amount of culturable virus pose the greatest degree of infectivity and risk to others. It should be stressed that viral loads for SARS-CoV-2 vary over nine orders of magnitude. The difference between the lowest viral loads, where virus is consistently detected by culture (10^5^ copies/mL, Fig. 1, 3), and the highest observed viral loads (∼1 billion copies/mL) is at least four orders of magnitude. Furthermore, above the viral culture detection threshold, we found that the amount of viable virus in day 3 culture supernatants was roughly proportional to sample viral load determined by qPCR (Fig. 1), suggesting that the large range of viral loads determined by RT-qPCR corresponds to the range and degree of sample infectivity. Importantly, antigen test sensitivity is noted to be near 100% in the upper three orders of magnitude of viral loads observed (Fig. 2, 3, 4), suggesting that antigen tests are quite good in detecting individuals who shed larger amounts of virions and therefore would pose significant risk to others during casual contact.

**Figure 4.**
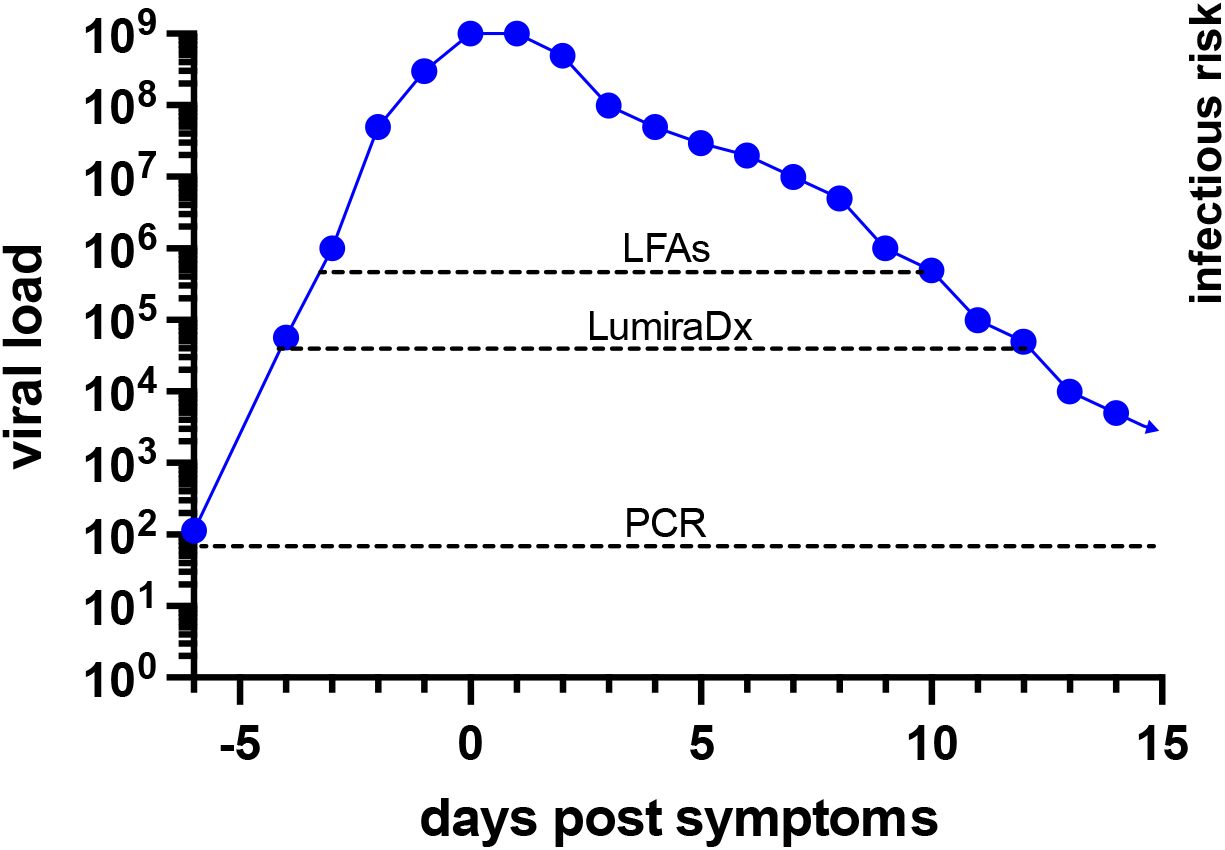
Model of Infectious Risk versus SARS-CoV-2 Detection by RT-qPCR and Antigen Tests. Both LumiraDx and lateral flow-based antigen tests (e.g., BD Veritor, CareStart, and Oscar Corona) were able to detect individuals with viable, culturable virus and who therefore pose an immediate infectious risk to others. Dotted lines indicate reliable detection threshold predicted for each method. Presumptively, infectious risk is proportional to the amount of culturable virus which is roughly proportional to the viral load in samples. Antigen tests were excellent in detecting patients with the highest viral loads which may be four to five log10-fold greater than viral loads detected at the lowest levels where virus can be consistently cultured. PCR and, to a lesser extent, the LumiraDx test can detect individuals before and after the expected infectious period and therefore may be more appropriate for screening programs where regular testing is performed at longer time intervals. The viral load curve shown is for representational purposes and may not reflect viral load kinetics in any specific individual.

It has been argued by some that Ct values should not be made available to providers based on a number of reasons, including differences in the correlation between Ct and viral load generated from different assays (reviewed in (13)). Our data support the meaningful association of viral load with infectivity, and therefore further argue for conversion of ambiguous Ct values to unambiguous viral loads, calibrated to a universal standard, as we have proposed previously (3).

Interestingly, the viral load threshold associated with consistent LFA detection in our clinical samples was similar to the previously described analytical limit of detection (LoD) of the Quidel Sofia antigen test determined using SARS-CoV-2 infected Vero cell quality control material (14). The LoD for the Sofia test in viral genome copies/mL was inferred from the TCID_50_ provided by the manufacturer in its Instructions for Use (IFU) (14), and the conversion factor between TCID_50_ and viral genome copies/mL provided in documentation for the lots (70033548, 70034991) of BEI Resources, NR-52286 quality control material available during the time the IFU studies were performed (3, 14). This correspondence suggests that quantitative relationships between detectable antigen and viral genomic material are similar for human nasopharyngeal specimens and virus grown in cell culture.

Our study design had several strengths as well as some limitations. It is known that there may be variability in results obtained from repeat sampling of the same patient, for example, due to collection technique (3, 15). We sought to eliminate this source of confounding variability by performing antigen, RT-qPCR, and viral culture testing on the same samples. Therefore, the per sample performance characteristics of each methodology could be directly compared.

However, this led to the need to perform antigen testing outside of direct swab sampling testing recommendations, potentially leading to underestimation of antigen test sensitivity. Based on the dilution factor in viral transport medium, and the amount of sample used in antigen testing in our study, we estimate that antigen tests, had they been performed by directly eluting swab samples into antigen test extraction buffer, would have detected samples with approximately 17 to 18-fold lower viral loads. As such, the antigen tests would have identified samples from individuals with the lowest viral loads associated with a positive viral culture. For example, we estimate that if swab samples were tested directly without dilution in viral transport medium, LFA antigen tests would have reliably detected individuals with viral loads > ∼550,000 genome copies/mL, and the LumiraDx test would have reliably detected individuals with viral loads > ∼60,000 copies/mL (see Fig 2). It is possible though that this estimate is too high or too low based on pre-analytical (e.g., sample elution efficiency) and analytical variables not appreciated.

Both a strength and weakness of the approach was the selection of samples based on the distribution of viral loads observed during clinical testing (3). Samples were not selected based on patients’ clinical symptoms, timing of symptoms, and potential exposures. Our testing centers obtain samples from community and hospital settings, both for diagnosis of illness and screening purposes. Thus, this is likely to reflect a real-world situation in which testing is performed for a variety of purposes. Therefore, without knowledge of the purpose and timing of specimen collection, it is possible that results may not be fully representative of performance characteristics in all diagnostic settings. Also, samples were obtained in March through June 2021 prior to emergence of the Delta or Omicron variants in our region. Relationships between viral load, antigen, and culture results are likely to be maintained. However, experimental verification during emergence of current and future variants would be desirable. Lastly, we used nasopharyngeal swab samples rather than nasal swab samples that are more often used for antigen testing assays. Of note, we previously found in a clinical trial, in which nasal swabs and nasopharyngeal swabs were collected from the same patients in parallel, that viral loads between these sample types were congruent for samples with viral loads > 1000 genome copies/mL (15), the range of interest in our study. Therefore, we believe that the use of nasopharyngeal swabs necessitated by our study design had negligible impact on our comparisons.

One goal was to determine whether gaps in specificity for each antigen testing method (i.e., false positive results) would or would not overlap. If the latter were found, then antigen tests could potentially be used sequentially for screening and confirmatory analysis. However, no false positives were noted for any method. Therefore, the number of samples analyzed was insufficient to address whether further improvements in aggregate specificity could be achieved by use of tandem antigen testing strategies. It is possible that our sampling strategy, resulting in specimen dilution compared with direct nasal sampling, may have been biased towards increased specificity.

Three of the antigen assays analyzed in our study (BD Veritor, CareStart, and Oscar Corona) were chromatographic LFAs. The BD Veritor system interprets results using an automated reader, while the CareStart and Oscar Corona assays was read visually. The Oscar Corona assay is not approved for use in the United States, but is widely used in India. The performance of these three assays in our study was essentially identical. In contrast, the LumiraDx assay used a microfluidic immunofluorescent detection technology and appeared significantly more sensitive than the other antigen test methods (see Fig. 1-3, Table 1), consistent with high level detection of infected patients during the first twelve days of symptom onset, as previously described, using PCR as the gold standard for infection (16). The observed lower sensitivity of antigen tests relative to PCR has also been described previously (17-20).

Informed use of SARS-CoV-2 testing is crucial to current and future control of the SARS-CoV-2 pandemic. The available testing options have different attributes in terms of TAT, potential for point-of-care deployment, cost, and performance. Our study finds that inexpensive, point-of-care deployable LFA assays have sufficient sensitivity to detect individuals whose diagnostic samples contain culturable virus and who therefore pose a potential transmission risk to others (Fig. 4). The performance characteristics of these LFA tests are outstanding in identifying the most highly infectious specimens. This attribute may be especially important during the SARS-CoV-2 Delta variant surge, characterized by individuals with viral load skewed to even higher levels, whether symptomatic or asymptomatic (21, 22).

Therefore, our data support use of antigen testing to identify infectious individuals at the time of sampling. These tests would presumably be highly efficacious at identifying and allowing isolation of significantly infectious individuals from communal events, same-day healthcare procedures, communal travel arrangements, and other settings with significant person-to-person contact where universal masking is not feasible or desired, as has been demonstrated in recent epidemiological studies (23). As a point-of-use testing modality, results can be immediately available and inform timely mitigation of infectious risk to others and/or clinical management. However, the sensitivity of antigen tests is low relative to PCR. They, therefore, will not identify recently infected individuals whose viral load and infectious burden has yet to climb into a detectable range, nor identify patients who have been infected in the recent past and whose results may inform contact tracing efforts. Accordingly, antigen tests lack the power of PCR for screening programs intended to secure populations through regular testing at longer spaced time intervals and testing of patients being admitted to hospitals where the best available analytical sensitivity is desirable to prevent outbreaks in at risk populations. The improved detection by the LumiraDx test shows that new testing modalities should be evaluated on a sliding scale relative to a quantitative standard such as viral load, for as in this case, they may provide an enhanced safety zone for screening individuals who will have contact with vulnerable populations or healthcare settings.

The identification of individuals at high risk for transmitting SARS-CoV-2 is a major public health goal to limit community spread. While viral culture may be the standard diagnostic method to determine infectivity; cost, complexity, and BL3 requirements prohibit its use as a routine clinical diagnostic method. Overall, our study supports use of antigen detection tests for specific purposes, where immediate detection of potential infectivity and especially highly infectious individuals is desired. Furthermore, although significantly less sensitive than PCR for detecting infected individuals, they provide a point-of-use alternative which may, through repeated testing at closely spaced time intervals and more rapid results, provide equivalent power to address goals of the pandemic at much lower cost.

## Materials and Methods

### Samples

The 206 SARS-CoV-2 positive and 300 SARS-CoV-2 negative samples analyzed in this study were nasopharyngeal swabs obtained in 3 mL of saline or viral transport medium at COVID-19 testing sites at Beth Israel Deaconess Medical Center (Boston, MA) for purposes of diagnosis unrelated this study. Samples were collected from March 2021 through June 2021, and selected for analysis solely based on viral load distribution. Specimens generally were collected at drive-through testing sites in Boston and several surrounding communities affiliated with our medical center (15). After PCR testing for clinical purposes, samples were stored at 4°C until testing by viral culture and antigen testing. Human subjects research in this study was approved by the Institutional Review Boards at Beth Israel Deaconess Medical Center and the Harvard T.H. Chan School of Public Health.

### RT-qPCR testing

SARS-CoV-2 RT-qPCR testing of samples and Vero cell culture supernatants was performed using the Abbott Molecular M2000 Real-Time or Alinity m SARS-CoV-2 assays according to the manufacturer’s instructions. Both assays had received Emergency Use Authorization (EUA) for qualitative diagnosis of SARS-CoV-2 infection and detect identical SARS-CoV-2 N and RdRp gene targets. In addition, they both output a quantitative fractional cycle number (FCN), a type of cycle threshold described in detail elsewhere (24). An extended panel of standards ranging from 300 to 10^6^ viral genome copies/mL (provided by LGC Seracare, Milford, MA) was used to establish a calibration curve and convert FCN values to viral genome copies/mL. The standards consist of replication-incompetent, enveloped, positive singled-stranded RNA Sindbis virus into which the whole genome of SARS-CoV-2 was cloned and titers determined using digital droplet PCR analysis by LGC SeraCare (Russell Garlick, LGC SeraCare, personal communication). The standards therefore model SARS-CoV-2 virus and were run through all stages of sample preparation and extraction to allow appropriate comparison with identically processed patient samples. Coefficients of determination (R^2^) for comparison of FCN values with log10 transformed viral load values obtained from analysis of standards were 0.997 for both assays. Slope and intercepts defined linear regression equations that were used to convert FCN to viral load values including extension above and below the level of calibrators tested. Evaluation of the accuracy and modeling of viral load conversions were described previously (3, 25).

### Antigen testing

The BD Veritor (Franklin Lakes, NJ), LumiraDx (Waltham, MA), CareStart (Access Bio, Inc., Somerset, NJ), and Oscar Corona (Oscar Medicare Pvt. Ltd, New Delhi, India) SARS-CoV-2 antigen tests were performed according to the manufacturer’s instructions with the exception that 250 uL of patient sample (nasopharyngeal swab sample eluted into 3 mL of saline or viral transport medium) was pipetted into the extraction vial provided with each kit rather than direct insertion of the nasal swab into the extraction vial. The LumiraDx test contained ∼600 uL of extraction buffer; the other methods used ∼500 uL of extraction reagent. Therefore, approximate dilution of sample compared with direct sample assuming complete elution from direct swab sampling was ∼17 fold for the Lumira method and ∼18-fold for the other antigen test methods.

### SARS-CoV-2 viral culture

Vero E6 (ATCC CRL-1586) cells were seeded on a 6-well flat bottom plate at 0.3 × 10^6^ cells per well in Eagle’s minimum essential media (EMEM) containing 1% antibiotic-antimycotic, 1% HEPES and 5% fetal calf serum (FCS, Gibco), and grown to confluence at approximately 1 × 10^6^ cells per well (9-11).

Vero E6 cells were inoculated with 250ul of patient sample and incubated at 37°C for 24 hours for viral adsorption. Simultaneously, a negative control was also inoculated with 250uL of viral growth media. Carryover of non-viable viral RNA present in samples was limited by washing cell cultures after the 24-hour viral adsorption and adding fresh EMEM composite media with reduced FCS to 2% for viral growth. Therefore, detectable virus should represent viable replicating virus. On days, 3, 6, and 13-14 days of culture, 800 uL of cell culture supernatant was removed and added to 800 ul of VXL buffer (QIAGEN, German, MD) (1:1 ratio) for subsequent nucleic acid extraction and SARS-CoV-2 real-time RT-qPCR. Cultures were re-fed with addition of 1 mL of EMEM with reduced FCS after sampling at each time point.

Supernatant viral loads below the LoD of the PCR assay were scored negative at that timepoint. Samples with two of three sequential supernatant viral loads exceeding the LoD were considered positive, and, conversely, samples with either one or no viral loads exceeding the LoD were considered negative.

### Statistics

Statistical comparisons were performed with Stata version 13.1 (Stata Corporation, College Station, TX) and/or Prism 9 for MacOS (GraphPad, San Diego, CA). Sensitivity and specificity for ROC curve analysis was determined through standard formulas in Microsoft Excel and imported into Prism for graphical representation.

## Data Availability

All data produced in the present work are contained in the manuscript

## Acknowledgements

This study was supported by an Accelerating Coronavirus Testing Solutions grant from the Massachusetts Life Sciences Center. We thank LumiraDx for providing instrumentation and antigen test kits; Oscar Medicare Pvt. Ltd for providing Oscar Corona antigen test kits; and Ginkgo Biosciences (Boston, MA) for providing the CareStart antigen test kits. We thank LGC SeraCare for providing reagents used in calibrating SARS-CoV-2 viral load assays. We received support from Abbott Molecular unrelated to this study under a COVID-19 Diagnostics Evaluation Agreement. One co-author, RA, was also a recipient of grant support from Abbott Molecular under a clinical study agreement. LumiraDx, Oscar Medicare Pvt. Ltd, LGC SeraCare, Abbott Molecular and Ginkgo Biosciences had no role in study design, manuscript preparation or decision to publish. All authors, no other conflicts of interest.

